# Comparative Household Secondary Attack Rates associated with B.1.1.7, B.1.351, and P.1 SARS-CoV-2 Variants

**DOI:** 10.1101/2021.06.03.21258302

**Authors:** Kevin A. Brown, Semra Tibebu, Nick Daneman, Kevin Schwartz, Michael Whelan, Sarah Buchan

## Abstract

**Background:** The emergence of SARS-CoV-2 variants associated with increased transmissibility are driving a 3^rd^ global surge in COVID-19 incidence. There are currently few reliable estimates for the P.1 and B.1.351 lineages. We sought to compare the secondary attack rates of SARS-COV-2 mutations and variants in Canada’s largest province of Ontario, using a previously validated household-based approach.

**Methods:** We identified individuals with confirmed SARS-CoV-2 infection in Ontario’s provincial reportable disease surveillance system. Cases were grouped into households based on reported residential address. Index cases had the earliest of symptom onset in the household. Household secondary attack rate was defined as the percentage of household contacts identified as secondary cases within 1-14 days after the index case.

**Results:** We identified 26,888 index household cases during the study period. Among these, 7,555 (28%) were wild-type, 17,058 (63%) were B.1.1.7, 1674 (6%) were B.1.351 or P.1, and 601 (2%) were non-VOC mutants (Table 1). The secondary attack rates, according to index case variant were as follows: 20.2% (wild-type), 25.1% (B.1.1.7), 27.2% (B.1.351 or P.1), and 23.3% (non-VOC mutants). In adjusted analyses, we found that B.1.1.7, B.1.351, and P.1 index cases had the highest transmissibility (presumptive B.1.1.7 OR_adjusted_=1.49, 95%CI 1.36, 1.64; presumptive B.1.351 or P.1 OR_adjusted_=1.60, 95%CI 1.37, 1.87).

**Discussion:** Substantially higher transmissibility associated with variants will make control of SARS-CoV-2 more difficult, reinforcing the urgent need to increase vaccination rates globally.

## Background

The emergence of SARS-CoV-2 variants associated with increased transmissibility are driving a 3^rd^ global surge in COVID-19 incidence. Two mutations, N501Y and E484K, are thought to be responsible for increased transmissibility of the B.1.1.7 (N501Y alone) and P.1/B.1.351 (N501Y and E484K) variants. Over the first months of 2021, the B.1.1.7 variant of concern (VOC) rapidly overtook wild type strains in the United Kingdom and many other jurisdictions, including Canada, and has been estimated to be 1.3 to 1.5 times more transmissible that previously circulating variants.^1–3^ There are currently few reliable estimates for P.1 and B.1.351. We sought to compare the secondary attack rates of SARS-COV-2 mutations and variants in Canada’s largest province of Ontario, using a previously validated household-based approach.^4^

**Table 1.** Secondary attack rates of persons infected with SARS-CoV-2, March 1 to April 17.

|  | Wild-type<br>(N501Y- and E484K-) |  | Presumptive<br>B.1.1.7*<br>(N501Y+ and E484K-) |  | Presumptive<br>B.1.351 or P.1*<br>(N501Y+ and E484K+) |  | Non-VOC mutant<br>(N501Y- and E484K+) |  |
| --- | --- | --- | --- | --- | --- | --- | --- | --- |
|  | Index Cases<br>N (%) | SAR<br>(%) | Index Cases<br>N (%) | SAR<br>(%) | Index Cases<br>N (%) | SAR<br>(%) | Index Cases<br>N (%) | SAR<br>(%) |
| Total | 7555 | 20.2 | 17058 | 25.1 | 1674 | 27.2 | 601 | 23.3 |
| Gender** |  |  |  |  |  |  |  |  |
| Female | 3746 (49.6) | 19.5 | 8051 (47.2) | 24.3 | 780 (46.6) | 26.2 | 292 (48.6) | 25.1 |
| Male | 3770 (49.9) | 20.9 | 8944 (52.4) | 25.9 | 889 (53.1) | 28.0 | 308 (51.2) | 21.8 |
| Age (median, IQR) | 36 [24, 53] |  | 37 [25, 51] |  | 39 [26, 54] |  | 36 [25, 52] |  |
| Age |  |  |  |  |  |  |  |  |
| [0,10) | 366 (4.8) | 18.3 | 706 (4.1) | 27.6 | 57 (3.4) | 26.2 | 34 (5.7) | 21.3 |
| [10,20) | 865 (11.5) | 15.7 | 1676 (9.8) | 22.0 | 138 (8.2) | 22.0 | 41 (6.8) | 15.4 |
| [20,30) | 1587 (21.0) | 16.2 | 3838 (22.5) | 20.9 | 348 (20.8) | 24.4 | 146 (24.3) | 18.8 |
| [30,40) | 1356 (18.0) | 20.0 | 3153 (18.5) | 24.5 | 299 (17.9) | 25.9 | 101 (16.8) | 23.3 |
| [40,50) | 1138 (15.1) | 22.5 | 2864 (16.8) | 26.3 | 280 (16.7) | 27.4 | 99 (16.5) | 31.4 |
| [50,60) | 1071 (14.2) | 24.1 | 2635 (15.4) | 29.3 | 297 (17.7) | 30.0 | 86 (14.3) | 25.6 |
| [60,70) | 711 (9.4) | 25.4 | 1418 (8.3) | 29.5 | 156 (9.3) | 33.3 | 62 (10.3) | 22.7 |
| [70,80) | 306 (4.1) | 28.7 | 584 (3.4) | 31.3 | 72 (4.3) | 35.3 | 23 (3.8) | 26.2 |
| [80+] | 153 (2.0) | 26.5 | 184 (1.1) | 32.5 | 27 (1.6) | 37.0 | 9 (1.5) | 35.3 |
| Reported Week |  |  |  |  |  |  |  |  |
| 9 (Feb 26-Mar 4) | 666 (8.8) | 20.3 | 223 (1.3) | 32.1 | 23 (1.4) | 30 | 2 (0.3) | 87.5 |
| 10 (Mar 5- 11) | 1456 (19.3) | 21.4 | 319 (1.9) | 29.2 | 79 (4.7) | 30.1 | 4 (0.7) | 11.1 |
| 11 (Mar 12-18) | 1315 (17.4) | 20.9 | 511 (3.0) | 25.1 | 104 (6.2) | 34.7 | 5 (0.8) | 0.0 |
| 12 (Mar 19-25) | 1118 (14.8) | 20.4 | 1401 (8.2) | 26.8 | 184 (11.0) | 25.3 | 39 (6.5) | 29.6 |
| 13 (Mar 26-Apr 1) | 1143 (15.1) | 18.8 | 2851 (16.7) | 25.7 | 273 (16.3) | 29.1 | 104 (17.3) | 21.9 |
| 14 (Apr 2-8) | 941 (12.5) | 19.5 | 4393 (25.8) | 26.0 | 399 (23.8) | 30.4 | 128 (21.3) | 23.1 |
| 15 (Apr 9-15) | 764 (10.1) | 19.5 | 5644 (33.1) | 24.1 | 490 (29.3) | 23.4 | 236 (39.3) | 22.4 |
| 16 (Apr 16-22) | 152 (2.0) | 18.5 | 1716 (10.1) | 22.4 | 122 (7.3) | 21.8 | 83 (13.8) | 24.9 |
| Symptom onset to<br>test, delay*** |  |  |  |  |  |  |  |  |
| No symptoms | 681 (9.1) | 7.9 | 1001 (5.9) | 9.5 | 104 (6.3) | 8.3 | 43 (7.3) | 7.8 |
| Pre-symptomatic | 284 (3.8) | 9.7 | 596 (3.5) | 13.5 | 63 (3.8) | 11.0 | 19 (3.2) | 1.7 |
| 0 days | 689 (9.2) | 13.8 | 1524 (9.0) | 19.7 | 145 (8.8) | 22.6 | 56 (9.5) | 14.1 |
| 1 day | 1371 (18.4) | 18.2 | 2976 (17.7) | 24.9 | 305 (18.4) | 27.3 | 92 (15.6) | 27.0 |
| 2 days | 1353 (18.1) | 22.1 | 3190 (18.9) | 26.3 | 296 (17.9) | 30.8 | 111 (18.8) | 24.7 |
| 3 days | 942 (12.6) | 23.4 | 2459 (14.6) | 27.1 | 245 (14.8) | 28.1 | 89 (15.1) | 25.9 |
| 4 days | 659 (8.8) | 27.1 | 1646 (9.8) | 28.3 | 144 (8.7) | 29.5 | 61 (10.3) | 25.8 |
| 5+ days | 1485 (19.9) | 25.8 | 3466 (20.6) | 29.5 | 353 (21.3) | 31.9 | 120 (20.3) | 29.7 |

SAR: secondary attack rate; \* lineages inferred from mutations based on local epidemiology; \*\* does not include missing or other genders (108 index cases and 67 associated secondary cases and 294 household contacts); \*\*\* testing delay: days between the index case symptom onset and specimen collection. Pre-symptomatic index cases with a testing delay of <0 days. Those that did not report a symptom onset date, or did not report a specimen collection date, were excluded from this section (320 index cases and associated 697 secondary cases and 2,578 household contacts).

## Methods

We identified individuals with confirmed SARS-CoV-2 infection in Ontario’s provincial reportable disease surveillance system.^4^ Cases were grouped into households based on reported residential address.^4^ Index cases had the earliest of symptom onset in the household.^4^ We included households with index cases were reported from March 1 to April 17, 2021, a period when all specimens were eligible for N501Y and E484K mutation testing. Households with multiple cases with the same earliest symptom onset date, were excluded.

Index cases were classified as: (1) wild-type (N501Y- and E484K-; predominantly B.1.2 and B.1.438.1^5^), (2) presumptive B.1.1.7 (N501Y+ and E484K-), (3) presumptive B.1.351 or P.1 (N501Y+ and E484K+), or (4) Non-VOC mutants (N501Y- and E484K+; predominantly B.1.525, and B.1.1.318^5^). We also included analysis based on B.1.1.7, B.1.351, and P.1 index cases confirmed by whole genome sequencing. Household secondary attack rate was defined as the percentage of household contacts identified as secondary cases within 1-14 days after the index case.

Logistic count regression was used to calculate unadjusted and adjusted odds ratios (OR) and 95% confidence intervals (CI), with the count of secondary cases and non-cases as the outcome and mutation group as the predictor. Random intercepts were included to account for household and health region clustering. The adjusted model included index case characteristics (gender, age, reported date, days from symptom onset to testing), and neighbourhood characteristics from the 2016 Canadian Census (proportion of visible minority residents, and household crowding). Statistical analysis was performed in R (v4.0.4). We obtained ethics approval from Public Health Ontario’s Research Ethics Board.

## Results

We identified 26,888 index household cases during the study period. Among these, 7,555 (28%) were wild-type, 17,058 (63%) were B.1.1.7, 1674 (6%) were B.1.351 or P.1, and 601 (2%) were non-VOC mutants (Table 1). The secondary attack rates, according to index case variant were as follows: 20.2% (wild-type), 25.1% (B.1.1.7), 27.2% (B.1.351 or P.1), and 23.3% (non-VOC mutants).

In adjusted analyses (Table 2), we found that B.1.1.7, B.1.351, and P.1 index cases had the highest transmissibility (presumptive B.1.1.7 OR_adjusted_=1.49, 95%CI 1.36, 1.64; presumptive B.1.351 or P.1 OR_adjusted_=1.60, 95%CI 1.37, 1.87). Index cases that were non-VOC mutants also had a significantly increased relative transmissibility compared to wild-type (OR_adjusted_=1.32, 95%CI: 1.04, 1.69). We observed similar increased transmissibility when separating out confirmed B.1.351 (OR_adjusted_=1.58: 95%CI: 0.93, 2.67) and P.1 lineages (OR_adjusted_=1.62, 95%CI: 1.21, 2.16).

**Table 2.** Unadjusted and adjusted odds ratios for the secondary attack rate, March 1 to April 17.

| Mutation or Lineage Profile | Index Cases<br>(N) | Secondary<br>Attack Rate<br>(%) | Odds Ratio (95% CI) |  |
| --- | --- | --- | --- | --- |
|  |  |  | Unadjusted | Adjusted** |
| Wild-type (N501Y-/E484K-) | 7,555 | 20.2 | Reference | Reference |
| Presumptive B.1.1.7 (N501Y+/E484K-) | 17,058 | 25.1 | 1.31 (1.21, 1.42) | 1.49 (1.36, 1.64) |
| Confirmed B.1.17* | 617 | 24.3 | 1.30 (1.02, 1.64) | 1.63 (1.28, 2.08) |
| Presumptive B.1.351 or P.1 (N501Y+/E484+) | 1,674 | 27.2 | 1.54 (1.33, 1.78) | 1.60 (1.37, 1.87) |
| Confirmed P.1* | 340 | 28.2 | 1.65 (1.24, 2.20) | 1.62 (1.21, 2.16) |
| Confirmed B.1.351* | 96 | 26.4 | 1.75 (1.03, 2.95) | 1.58 (0.93, 2.67) |
| Non-VOC mutant (N501Y-/E484K+) | 601 | 23.3 | 1.19 (0.94, 1.50) | 1.32 (1.04, 1.69) |

VOC=variant of concern.
\* Subset of samples confirmed with whole genome sequencing;
\*\* Adjustment covariates included index case gender, age, reported date, the number of days between symptom onset and test date, and neighbourhood proportion of visible minority residents (non-white and non-Indigenous population) and household crowding

## Discussion

In this study including over 20,000 households, we observed increased transmissibility from index cases with the N501Y mutation (B.1.1.7), both N501Y and E484K mutations (B.1.351 and P.1), and to a lesser extent, the E484K mutation alone. Limitations include lacking data on vaccination, and potential index case misclassification. Although we inferred variants from their mutation profile, results among cases confirmed by whole genome sequencing were consistent. Substantially higher transmissibility associated with variants will make control of SARS-CoV-2 more difficult, reinforcing the urgent need to increase vaccination rates globally.

## Data Availability

The specific dataset used for this analysis is not publically available. Ontario's Ministry of Health does make a modified version of this data publicly available (https://covid-19.ontario.ca/data). Questions regarding access to data should be directed to the ministry of health through the above website.

## Notes

### Competing Interest Statement

The authors have declared no competing interest.

### Funding Statement

No sources of external funding were used for this research.

### Author Declarations

Statistical analysis was performed in R (v4.0.4). We obtained ethics approval from Public Health Ontario's Research Ethics Board.

## References

1. Volz E, Mishra S, Chand M, et al. Transmission of SARS-CoV-2 Lineage B.1.1.7 in England: Insights from linking epidemiological and genetic data. medrxiv. Published online January 4, 2021. doi:10.1101/2020.12.30.20249034

2. Brown KA, Gubbay J, Hopkins J, et al. S-Gene Target Failure as a Marker of Variant B.1.1.7 Among SARS-CoV-2 Isolates in the Greater Toronto Area, December 2020 to March 2021. JAMA. 2021;325(20):2115. doi:10.1001/jama.2021.5607

3. Buchan SA, Tibebu S, Daneman N, et al. Increased household secondary attacks rates with Variant of Concern SARS-CoV-2 index cases. medrxiv. Published online April 5, 2021. doi:10.1101/2021.03.31.21254502

4. Paul LA, Daneman N, Brown KA, et al. Characteristics associated with household transmission of SARS-CoV-2 in Ontario, Canada: A cohort study. Clinical Infectious Diseases. Published online March 5, 2021:ciab186. doi:10.1093/cid/ciab186

5. Phylogenetic Analysis of SARS-CoV-2 in Ontario. Public Health Ontario https://nextstrain.publichealthontario.ca/ncov

